# Treatment-related Cognitive Impairment, Neurovascular Coupling, and Iron Levels in Women with Ovarian Cancer

**DOI:** 10.64898/2026.07.22.26358743

**Authors:** Summer Edwards, Quinn Smith, Fan Zhang, Anna Kuan-Celarier, Lei Ding, Doris M Benbrook, Joan Walker, Dee H. Wu, Michael Wenger, Han Yuan

## Abstract

Chemotherapy is a common treatment for women with ovarian cancer, and many women treated with chemotherapy experience cognitive dysfunction. However, few studies have investigated the mechanisms of chemotherapy-related cognitive impairment in women with ovarian cancer. The goal of this study was to assess the relationships among neurovascular coupling, iron levels and cognitive functions in women with ovarian cancer after first-line chemotherapy. Simultaneous fNIRS and EEG data were collected in women diagnosed with advanced stage ovarian cancer at baseline (N = 13) and after 3-9 rounds of chemotherapy (N = 8). The attentional network task (ANT) was administered for neurocognitive evaluation. Blood iron levels were measured using standard clinical assays. In parallel, similar concurrent fNIRS-EEG data were acquired in a group of 10 healthy participants in a test-retest design. Cognitive performance declined, as indicated by decreases in ANT sub scores after chemotherapy. Blood iron biomarkers indexing oxygen transport also declined and were related to decreases in the ANT sub scores. Both fNIRS and EEG data responses were shown to have excellent reliability in the healthy subjects, while cancer patients showed significant decreases in oxygenated hemoglobin response in fNIRS, despite no changes in EEG responses after chemotherapy. A significant dose relationship was also found in the changes of fNIRS responses. Our data indicate that chemotherapy produced cognitive deficits and decreases in the oxygenated hemoglobin response and that these may be related to reduced oxygen transport capacity. Results also suggest the utility of using fNIRS-EEG to monitor the progression of cognitive impairment and characterize those mechanisms.

## Introduction

It is estimated that ovarian cancer affects 1 in 87 women in their lifetime [1], with a combination of chemotherapy and debulking surgery as standard treatment. Considering that more than 50% of women with ovarian cancer survive at least five years [1], cognitive dysfunction is of significant concern as it may interfere with quality of life [2, 3]. Chemotherapy-related cognitive impairment (CRCI) in cancer survivors has been recognized as a serious issue in recent years and the mechanisms underlying CRCI are poorly understood. This poses serious challenges to developing improvements to managing CRCI during cancer treatment and managing chemotherapy related cognitive side effects [4].

Standard chemotherapy treatment results in high reports of “brain fog”, sometimes referred to as “chemo brain”. Up to 75% of patients receiving chemotherapy for any cancer report chemo brain, with many (up to 35%) continuing to report chronic symptoms after treatment concludes [5, 6]. CRCI symptoms include both general (e.g., subjective slowness and “fogginess”) and specific (problems with attention and memory) complaints. Recent research has characterized and assessed chemotherapy-related cognitive impairment in a number of specific cognitive domains, including, attention, memory, learning, language, executive function etc. [2, 7–9]. While these studies described chemotherapy-induced changes in cognitive abilities in ovarian cancer or other cancer populations, little is known about the neural mechanisms of the impairment. Consideration of possible mechanisms is complicated by the fact that many chemotaxic agents do not cross the blood-brain barrier [10].

One possible set of mechanisms may be related to the iron deficiency (ID) and iron deficiency anemia (IDA) induced by chemotherapy. Anemia is a highly prevalent consequence of cancer treatment [11–13]. A review by Xu et al. [14] found that, across all cancer types, 89.5% of patients developed some form of anemia. The lowest incidence was 86.3% for patients treated for breast cancer, and the highest incidence was 98.4% for patients treated for ovarian cancer. Patients treated for ovarian cancer had the highest prevalence (16.9%) of the severest form of anemia, defined as hemoglobin (Hb) levels ≤ 8 g/dL. A more recent review [15] found that women treated for ovarian and other gynecological cancers showed a > 240% increase in the odds of developing anemia and that < 25% of those women received treatment for the anemia. It is known that cytotoxic chemotherapy treatments induce anemia, impairing the synthesis of red blood cell precursors in bone marrow, decreasing the renal production of erythropoietin, or some combination of the two [15, 16]. The negative effects of ID/IDA include reductions in both Hb and sFt, and associated impairments in physical performance and work productivity [17–19], brain function [20–26], and cognition [27–30]. Especially, the impacts of ID/IDA on brain function and structure include effects on the synthesis, transport, and re-uptake of at least three of the catecholamine neurotransmitters— dopamine, norepinephrine, and serotonin—the transport of oxygen necessary for energy metabolism within the brain, and the production of myelin [31, 32]. All of these could play a role in the etiology and progression of CRCI and the subjective complaints regarding CRCI [33].

Previous studies to understand CRCI using various neuroimaging modalities have shown changes in the structure and function of the brain [2]. Structural studies using MRI and diffusion tensor imaging (DTI) have revealed reduced gray matter and white matter volumes as well as white matter integrity in the cortical and subcortical brain regions following chemotherapy [34–40]. Functional neuroimaging studies using task-based fMRI have shown involvement of cortical and subcortical circuitry that correlated significantly with subjective [41] and objective decreases in functions [35]. fMRI studies have also demonstrated the reduced resting-state functional connectivity patterns within the brain wide circuitry that are associated with cancer and chemotherapy in patients [42, 43]. Additionally, a few studies based on positron emission tomography (PET) and single-photon emission computed tomography (SPECT) have shown abnormal cerebral blood flow and metabolism in multiple brain regions of breast cancer survivors treated with chemotherapy [43–45]. While these functional studies have documented many changes in the brain, the measures of blood flow or hemodynamics do not directly speak to neural activity, as the hemodynamic measures are proxies for neural activity. More insight can potentially be gained by pairing a technology capable of resolving brain oxygen consumption with high temporal resolution measurement of neural activity.

In this paper, multimodal neuroimaging of concurrent functional near-infrared spectroscopy and electroencephalography (fNIRS-EEG) is used *for the first time* on women with ovarian cancer to assess longitudinal chemotherapy related-effects in the brain. This imaging system was used to record neural signals as well as behavioral responses at baseline and then after chemotherapy. fNIRS is a noninvasive and portable neuroimaging technique that uses near-infrared light to measure oxygenated and deoxygenated hemoglobin concentration changes via pairs of sources and detectors. Near-infrared light penetrates the scalp and is absorbed by oxygenated hemoglobin (HbO) and deoxygenated hemoglobin (HbR), while detectors measure the remaining light in a multiplexed manner [46–49]. In a healthy brain, increased neuronal activities typically leads to increased blood flow, which is referred to as neurovascular coupling (NVC) and is reflected as an inverse relationship between HbO and HbR [50]; specifically, as HbO increases, HbR decreases. However, the coupling process may be impaired in neurological diseases. For example, with age, the brain’s ability to effectively increase blood flow in response to increased neural activity declines and this impairment can contribute to cognitive decline and neurodegenerative disease [51]. However, in cancer patients it is not clear whether or how the neural activity and blood flow would react differently after chemotherapy. Therefore, here we used simultaneous EEG and fNIRS recording to examine the neurovascular process. EEG is a noninvasive tool that measures neuronal electrical activity via electrodes on the scalp. Neural activity can then be quantified as event-related potentials (ERPs), averaging electrical response from time-locked, stimulus blocks together. The ERP has been well established as the neuronal responses to tasks and linked to the hemodynamic measures [52]. Since fNIRS measures very similar signal changes to fMRI and is compatible with EEG recording, and since both technologies are relatively portable, the multimodal fNIRS and EEG can be feasibly set up in clinics to investigate the neurovascular coupling process in the ovarian cancer patients.

Previous use of simultaneous fNIRS-EEG has been done with human subjects across a range of tasks and conditions [47, 53, 54]. Considering that fNIRS and EEG measures in the primary motor cortex have been well characterized [54–56], our current study probed the neurovascular coupling in the motor cortex. We hypothesized that chemotherapy related NVC impairment may be present and could be measurable at the motor cortex where fNIRS and EEG responses are reliable. We recruited and studied 13 women diagnosed with advanced stage ovarian or endometrial cancer at baseline (BL) and after their course of chemotherapy (endline, EL). Participants provided blood samples that allowed us to quantify iron status before and after chemotherapy. Simultaneous fNIRS and EEG data were collected in the cancer patients during their clinic visits. Patients performed a hand clenching task for the neurovascular coupling measures as well as the attentional network task (ANT) [57, 58] for neurocognitive evaluation. In parallel, similar concurrent fNIRS-EEG data were acquired in a group of 10 healthy participants in a test-retest design, for the purpose of examining the reliability of fNIRS and EEG particularly in the concurrent setting. Consistent with the design of the ANT as a reaction time (RT) task, we quantified performance before and after chemotherapy in terms of three difference scores: one quantifying the extent to which attention is captured by low-level onsets of energy (referred to as altering), one quantifying the extent to which spatial cues are capable of being used to direct attention in space (referred to as orienting), and one quantifying the extent to which an individual can benefit from consistency and avoid costs due to inconsistency in attentional cues (referred to as conflict). We quantified NVC in terms of fNIRS and EEG responses in cancer patients and healthy subjects. In all these measures, we assessed whether there were any differences between the two repeated visits, which would be indicative of the chemotherapy treatment effect in the cancer patients.

## Results

### Demographic and Clinic Data

A total of 13 cancer patient participants meeting the inclusion/exclusion criteria were recruited and completed the study protocol from 2021 to 2024 (13 females, 38-85 years of age). While 13 patients were enrolled, only 8 patients who completed both visits are included in data analysis. The demographics and clinical characteristics of the cancer patients are listed in **Table 1**. Four of the eight patients had their baseline recording before they started chemotherapy, while four patients’ baseline recording occurred after the initial round of chemotherapy and before their second round (9.00 ± 8.36 days, range 2-21 days). At the second visit, all patients had completed 2-6 rounds of chemotherapy. In parallel to the patient study, a total of 10 healthy participants were recruited (4 females, 19-23 years of age) to assess the test-retest reliability of the NVC measures. The healthy subjects’ demographics are listed in the **Supplemental Table 1**.

**Table 1:** Cancer Subject Demographics.

| Subject (N = 8) | Age (Years) | Gender (F/M/Unknown) | Days Between Visit | Rounds of Treatment Between Visits |
| --- | --- | --- | --- | --- |
| Mean | 60.5 | 8/0/0 | 94.0 | 4 |
| STDV | 16.3 | - | 37.8 | 1.27 |

### Blood Assays

The results of the blood assays are presented in **Table 2** for the six participants for whom both baseline (BL) and endline (EL) assay results were available. Changes between EL and BL were assessed using one-sample two-sided *t*-test. Given the evidence for inflammation (C-reactive protein (CRP) > 5.0), serum ferritin (sFt) values were adjusted using the CRP values [63]. Values of hemoglobin (Hb), red blood cell count (RBC), and hematocrit (Hct) significantly decreased and red blood cell distribution width (RDW) significantly increased following chemotherapy, while adjusted sFt did not significantly change. The decrease in the iron biomarkers associated with oxygen transport, and the increase in RDW, in the context of high levels of adjusted sFt are consistent with a condition known as *functional* iron deficiency, in which iron stores are sufficient but are sequestered (due in part to inflammation) and unavailable to tissues and bone marrow [64, 65]. This would be consistent with other work documenting functional iron deficiency following chemotherapy [65, 66].

**Table 2:** Baseline (BL) and endline (EL) values of the blood assays for participants (*N* = 6 for whom both sets of assays were available).

| Variable | Baseline |  | Endline |  | Change | *t* | *p* |
| --- | --- | --- | --- | --- | --- | --- | --- |
|  | Mean | STDV | Mean | STDV |  |  |  |
| CRP, mg/L | 83.83 | 64.96 | 8.00 | 5.80 | -75.83 | 2.36 | 0.076 |
| **Hb, g/dL** | **12.50** | **1.94** | **10.57** | **1.16** | **-1.93** | **4.19** | **0.006** |
| **Hct, %** | **39.15** | **5.92** | **32.88** | **3.26** | **-6.27** | **3.98** | **0.008** |
| MCH, pg | 28.50 | 0.99 | 29.68 | 1.34 | 1.18 | 2.04 | 0.121 |
| MCHC, g/dL | 31.92 | 0.64 | 32.15 | 0.61 | 0.23 | 0.88 | 0.501 |
| MCV, fL | 89.28 | 1.39 | 92.40 | 5.02 | 3.12 | 1.36 | 0.300 |
| **RBC, M/mm<sup>3</sup>** | **4.38** | **0.67** | **3.58** | **0.52** | **-0.80** | **8.68** | **< 0.001** |
| **RDW, %** | **13.25** | **1.21** | **19.32** | **4.18** | **6.07** | **3.10** | **0.027** |
| sFt (adjusted), ng/mL | 166.86 | 133.39 | 243.50 | 122.10 | 76.64 | 1.78 | 0.174 |

Note: CRP: C-reactive protein, Hb: hemoglobin, Hct: hematocrit, MCH: mean corpuscular
hemoglobin: Hb, MCHC: mean corpuscular hemoglobin concentration, MCV: mean corpuscular
volume, RBC: red blood cell count, RDW: red blood cell distribution width, sFt: serum ferritin. **Bold**
indicates significant changes after chemotherapy ( $p < 0.05$ ).

### ANT Performance

The values of the three ANT performance measures – alerting, orienting, and conflict – at BL and EL are presented in **Table 3** for the six participants for whom both BL and EL performance data and BL and EL blood data were available. Note that reductions in all three measures are indicative of impairment to attentional function. Significance of changes was assessed using one-tailed *t*-tests. As can be seen in the table, there were significant reductions in all three measures, indicating impairment to all three levels of attentional functioning as a consequence of chemotherapy.

**Table 3:**
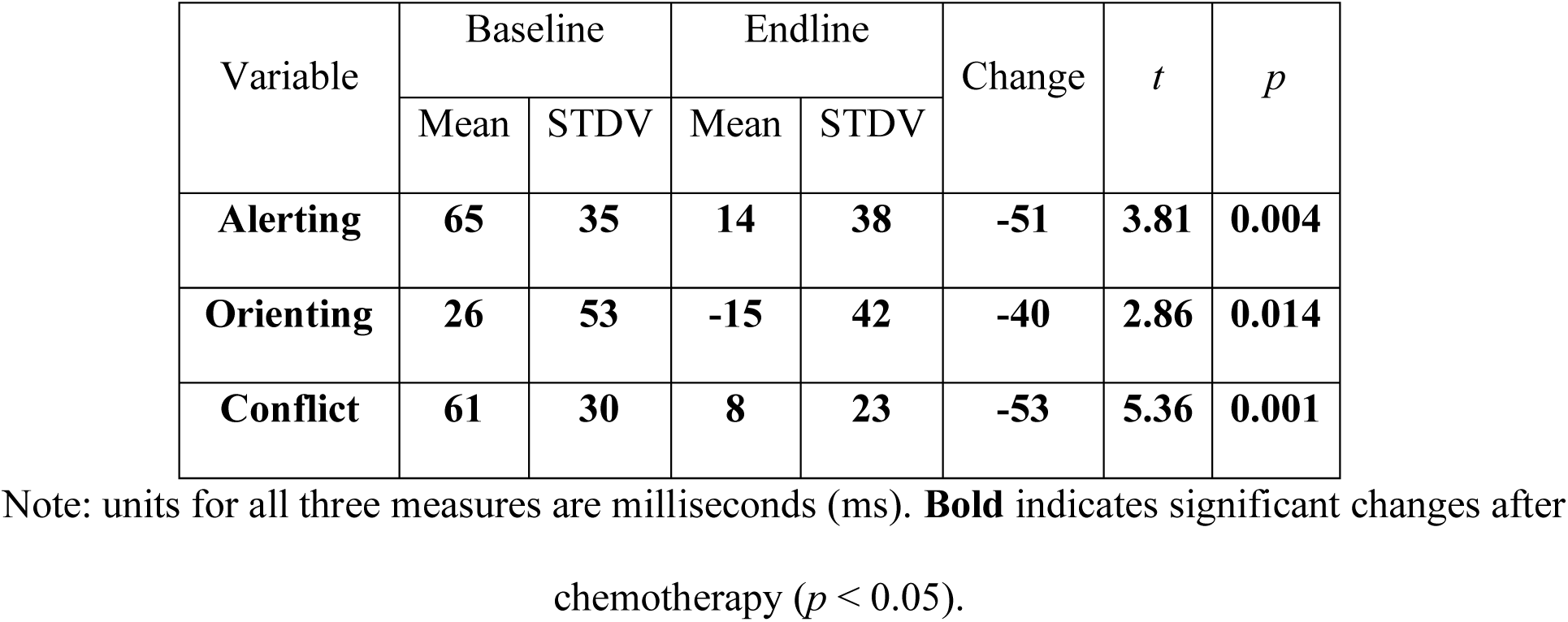
ANT performance measures (N = 6 for whom with complete behavioral and blood data).

### Neurovascular Coupling

At this point, we have established that, in a sample of women being treated with a chemotaxic agent that does not cross the blood-brain barrier, chemotherapy produces changes in iron status that are consistent with functional iron deficiency. Also, we have shown the decline of cognitive performance in these patients. We next consider impacts of chemotherapy on neurovascular coupling and tested whether there were differences in neurovascular coupling, specifically in the fNIRS and EEG responses, following chemotherapy. **Figure 1** shows data from a representative cancer patient at baseline and a representative healthy subject. In both subjects, the HbO activity shows a strong increase during the motor task and a return to baseline afterwards during the resting period. The spatial map of the activation indicates that the activation was focal over the left primary motor cortex. Similarly, in the concurrent EEG recordings, ERP from the recording channel over the motor cortex shows an initial slow and slight decrease and then followed by a strong increase. Comparing the data between the two repeated visits, the fNIRS and EEG responses from the healthy subject are very similar and their magnitude of reactions are consistent. However, there was a significant difference in the fNIRS data between visits in the cancer patient. The cancer patient’s HbO response was notably lower at EL relative to BL, although the ERP response did not show such a decrease after chemotherapy.

**Figure 1:**
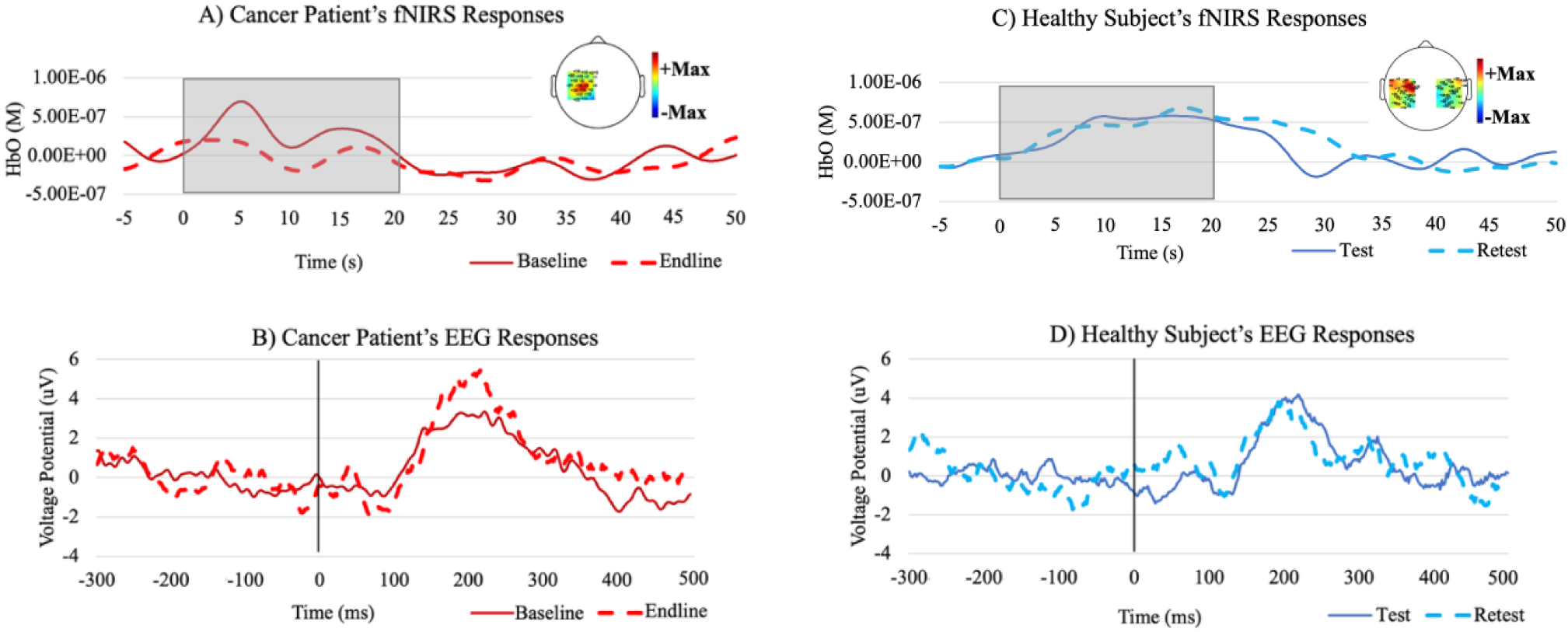
Neurovascular responses to the hand clenching task in a representative cancer patient and a healthy subject at two repeated visits. A) a cancer patient’s HbO response, B) a cancer patient’s EEG response, C) a healthy subject’s HbO response, and D) a healthy subject’s EEG response. The patient had two visits, one before and one before and after treatment (baseline and endline, respectively), while the healthy subject also had two repeated visit (test and retest, respectively). The inserts of topographic heat maps in (A) and (C) show fNIRS activity from all measurement channels with the peak activity over the left primary motor cortex. The grey shaded blocks in (A) and (C) indicate the task duration from 0 s to 20 s, and the black lines in (B) and (D) indicate the onset of each clenching at 0 ms.

We then quantified the neurovascular responses at the group level, with fNIRS and EEG responses. Only six cancer patients had complete EEG and fNIRS recordings at both BL and EL visits, while ten healthy subjects all had complete EEG and fNIRS at test and retest visits, respectively. First of all, performing the task has induced significant responses in fNIRS and EEG data from cancer and healthy subjects, which then allowed us to further compare the responses between visits. For the fNIRS (**Fig. 2)**, HbO signal increased as a reaction to the hand clenching task which is significant in both cancer patients and healthy subjects (healthy test, p = 0.004; healthy retest, p < 0.001; cancer BL, p = 0.003; cancer EL, p = 0.003). For the EEG (**Fig. 3**), ERP responses were also significant (all *p* < 0.001). Since both fNIRS and EEG measures significant neurovascular responses, we then examined the data between visits. In the heathy group, both fNIRS and EEG responses showed similar and consistent level of responses from the test and retest data (all *p* > 0.100). Additionally, the intraclass correlation coefficient (ICC) metric yielded 0.56 for fNIRS (good reliability) and 0.88 for EEG (excellent reliability). Meanwhile, in the cancer group, fNIRS response showed a significant reduction in the post-treatment relative to that at baseline (*p* = 0.028). Interestingly, however, there was no between-visit difference in EEG ERP responses for the cancer patients (*p* > 0.100). Although the healthy and cancer subjects were not matched in their age and gender, the cross-section comparison between the group groups yielded significance differences, with cancer patients showing lower HbO responses in the BL visit (*p* = 0.0483) and further lower HbO responses in the EL visit (*p* = 0.003). The patients’ EEG responses did not differ from those in healthy at either visit (all *p* > 0.100).

**Figure 2:**
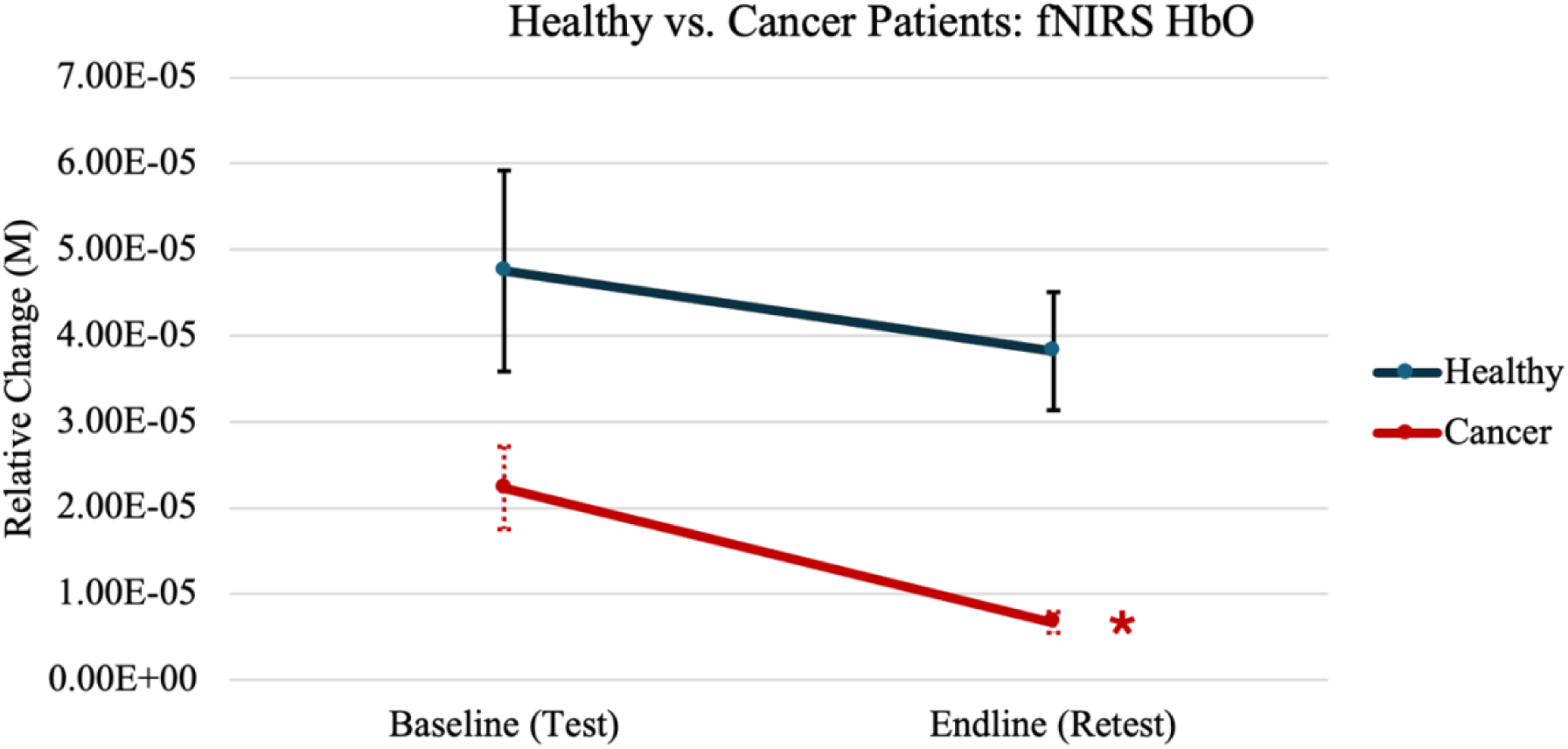
Group level comparison of fNIRS HbO response for the healthy and cancer patient groups. The patients had two visits at baseline and after treatment (baseline and endline, respectively), while the healthy subjects also had two repeated visit (test and retest, respectively). * indicates that the patients had a significant decrease (p = 0.028), but the healthy subjects did not show significant changes. Standard errors are drawn.

**Figure 3:**
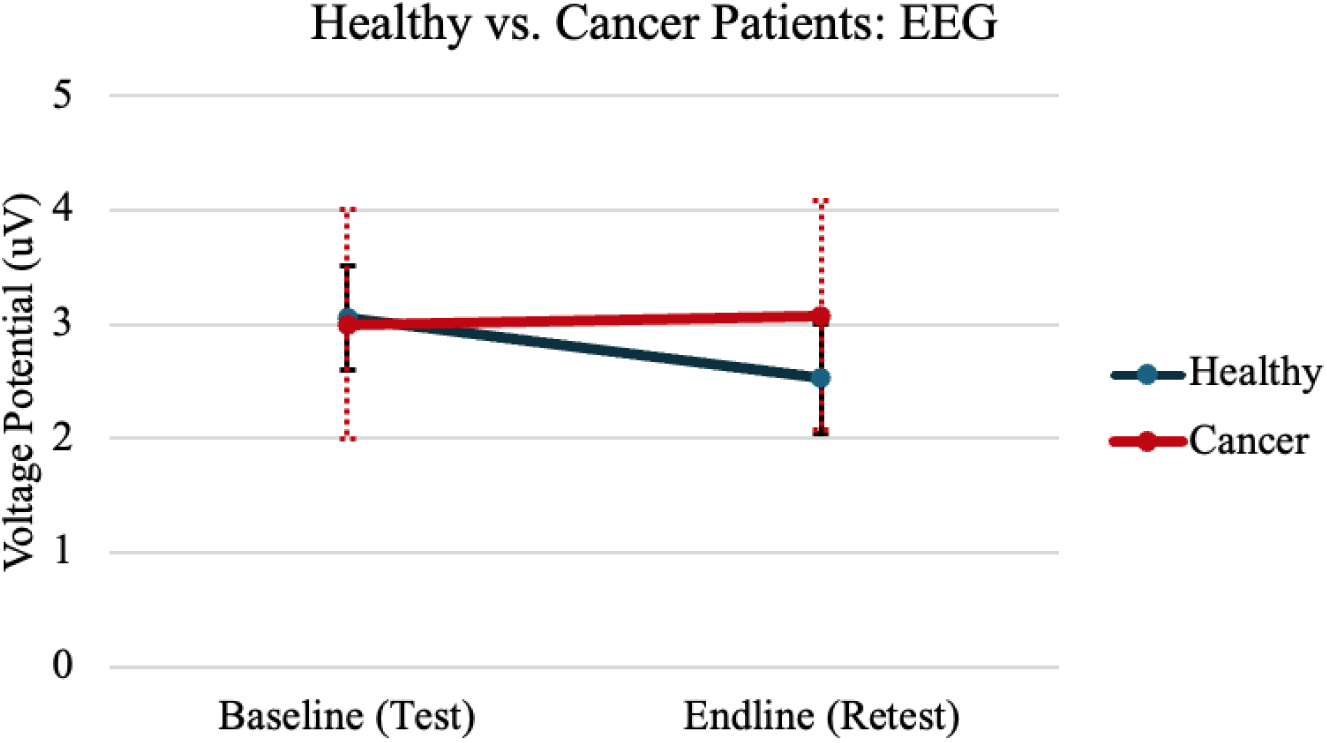
Group level comparison of peak amplitude of EEG event related potentials between healthy and cancer groups. The patients had two visits at baseline and after treatment (baseline and endline, respectively), while the healthy subjects also had two repeated visit (test and retest, respectively). There are no significant differences between visits for each group. Standard errors are drawn.

Next, we examined the dose relationship involving duration of chemotherapy, HbO, cognitive scores and blood iron biomarkers to investigate the extent to which the chemotherapy affects these measures. **Figure 4** shows a significant linear association for fNIRS response, such that more cycles of chemotherapy was related with a greater decrease in HbO (*r* = 0.90, *p* = 0.014). Also, we sought to determine whether there was any sign of a dose-response relationship between the number of rounds of chemotherapy and the level of impairment in attentional function. In all three cases, the dose effect shows a negative trend - more rounds of chemotherapy were associated with larger decrements in alerting, orienting and conflict functions, which however, did not reach significance (all *p* > 0.100). Finally, the dose effect on the hemoglobin in the blood count was negative but the linear association was not significant (all *p* > 0.100).

**Figure 4:**
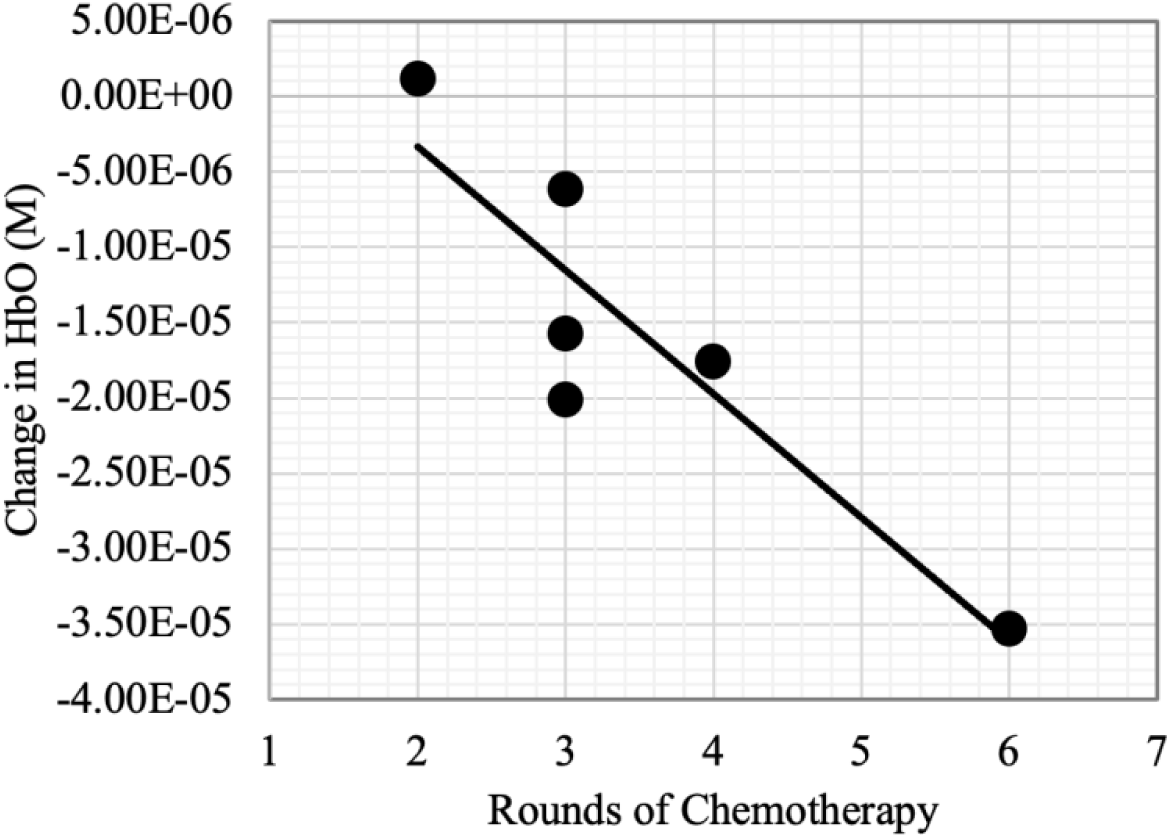
Dose relationship on fNIRS HbO change. Rounds of chemotherapy are associated with decreases in HbO response (*r* = 0.90, *p* = 0.0142).

Next, in order to understand the mechanism of these changes, we examined the data for the extent to which the declines in attentional performance or HbO were at all related to changes in the blood and ion biomarkers that showed significant changes from BL to EL – Hb, RBC, and Hct – along with the one biomarker suggestive of functional iron deficiency – RDW. Results revealed that changes indicative of reduced iron status (reductions in Hb, RBC, and Hct; and increases in RDW) were significantly related to decrements in attentional performance (**Fig. 5**). However, in the five subjects that had complete blood and fNIRS HbO measurements, only reductions in RBC were marginally related to changes in HbO (*r* = 0.84, *p* = 0.06), while none of the Hb, HCT or RDW were significantly related to HbO changes (all *p* > 0.1). There is a trend of positive association between HbO and cognitive functions, which however, did not reach significance (all *p* > 0.100). Functional iron deficiency following chemotherapy is a state in which iron stores (as measured by sFt) are sufficient but, due to a variety of factors, iron is not metabolically available to tissues and bone marrow. This would include availability for neurotransmitter synthesis and regulation, brain energy expenditure, and erythropoiesis. The combination of negative impacts on these processes could mechanistically underlie CRCI.

**Figure 5:**
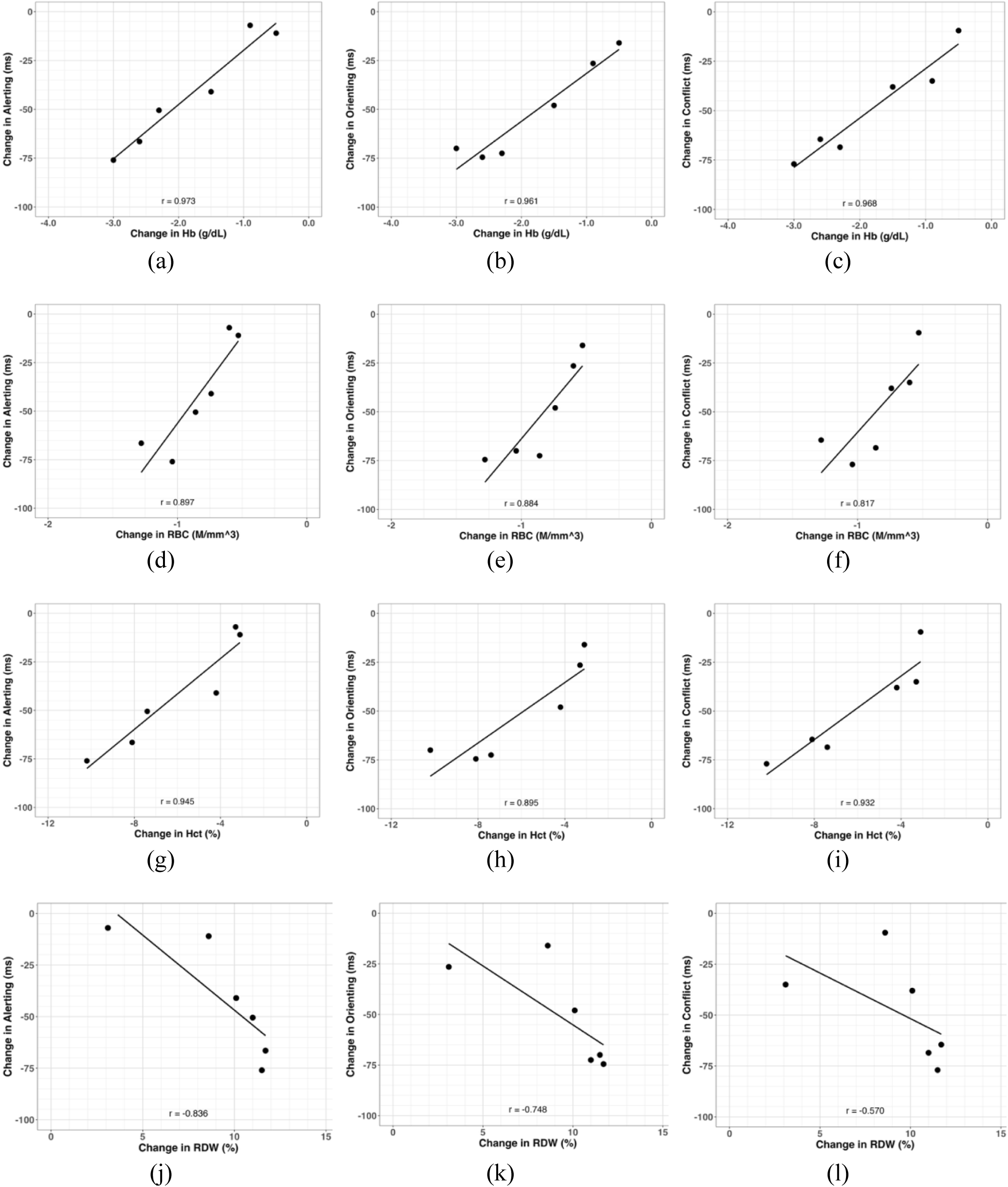
Relationships between changes in blood biomarkers and the three attention network task performance scores: (a)-(c) changes in hemoglobin (Hb) and changes in performance, (d)-(f) changes in red blood cell (RBC) and changes in performance, (g)-(i) changes in hematocrit (HCT) and changes in performance, (j)-(l) changes in red blood cell distribution width (RDW) and changes in performance. Note: Correlations in (a)-(j) are significant at *p* < 0.05, (k) is marginally significant (p = 0.087), but (l) did not reach significance (*p* > 0.100).

## Discussion

The current study used a unique multimodal imaging approach involving concurrent fNIRS and EEG to measure the neurovascular coupling process in ovarian cancer patients before and after chemotherapy treatment. Through multimodal measurements, we assessed and quantified the extent to which chemotherapy may induce changes in neurovascular coupling and the effects of chemotherapy on systemic iron levels. This allowed us to consider the multiple ways in which reductions in iron levels (and possibly functional iron deficiency) might be some of the mechanisms underlying CRCI: reductions in red blood cell formation, oxygen to the brain, oxygen transport within the cortex (as measured by HbO), along with impacts on neurotransmitter synthesis and regulation.

We first demonstrated that, consistent with the literature on chemotherapy, treatment resulted in significant reductions in a set of biomarkers related to oxygen transport and erythropoiesis (Hb, RBC, and HCT), while not affecting the biomarker indicative of iron storage (sFt). This combined pattern of results suggests that chemotherapy resulted in a state of functional iron deficiency, though additional assays would be needed to determine this. We then demonstrated that chemotherapy also resulted in decrements in three levels of attentional function and that reductions in measures of those three aspects of attention were systematically related to changes in the iron biomarkers, which is consistent with reports of CRCI in larger cohorts [59, 60].

Having established the negative impacts of chemotherapy on iron status and cognitive performance, we then demonstrated that, *for the first time*, that there was a significant reduction of the fNIRS HbO response but no change in the EEG response to the hand clenching task in cancer patients after chemotherapy. Critically, a dose analysis revealed that greater amount of chemotherapy is linearly associated with greater decrease of fNIRS HbO response. Noteworthy, the HbO reductions are associated with reductions in RBC with marginal significance. In terms of functional relevance, there is a trend of positive association between HbO responses and cognitive functions, which however, did not reach significance. We suggest that this is most likely due to the small sample size, a point to which we return below.

Our study is the first to report a longitudinal change in the neurovascular coupling using a unique multimodal approach of concurrent fNIRS and EEG. The CRCI studies so far have documented a number of changes to brain structure and function changes using various imaging modalities. Among these studies, the majority of them have examined the patients with breast or lung cancer. Few CRCI studies have considered patients with ovarian cancer, and those have taken a cross-sectional approach. For example, a structural imaging study of ovarian cancer patients using voxel-based morphometry showed that patients had significantly reduced gray matter volume compared to healthy controls in the right middle/superior frontal gyrus, and in the left supramarginal gyrus and left inferior parietal lobule [9], indicating abnormality in largely distributed brain areas that may be relevant to the negative effects of chemotherapy. Functional neuroimaging scans of ovarian cancer patients treated with chemotherapy have found lower resting metabolism [67], lower local synchronization of resting state fMRI [68], and reduced activation while performing memory tasks [9] in patients after chemotherapy compared to healthy controls. The abnormality of functional changes reported in these studies are spread out over the brain, affecting largely distributed areas [2]. To the best of our knowledge, our study reports, *for the first time*, a reduction of hemodynamic response after chemotherapy and a dose/response relationship with the chemotherapy. The area we chose to characterize for the neurovascular coupling process is the motor area, which has been found to present abnormally in prior structural and functional studies of cancer treatment [9, 67, 68]. Although it is not an area that directly underlies cognitive functions, it is involved in the generation of the motor response indicative of the completion of cognitive processing, and the reduction of response to the motor task is similar to that reduction seen in the response to working memory task [9] and is consistent with the network-level impairment in the brain [2].

Our finding of the reduction of the hemodynamic response after chemotherapy is related to other studies with cancer patients. For example, in a longitudinal study of patients with breast cancer, McDonald et al. documented that chemotherapy reduced fMRI activation during a working memory task, while in patients who did not receive chemotherapy there was no such reduction [69]. Additionally, Durán-Gómez et al. measured cerebral oximetry in patients with breast cancer as they performed a verbal fluency task and found that the patients’ responses to the task in the prefrontal cortex was reduced after chemotherapy [70]. In line with these two studies, we also found that the HbO response to the motor task decreased after chemotherapy, with this occurring in the context of reductions in systemic Hb, RBC, Hct and RDW. Furthermore, the available data indicate that storage iron, as measured by sFt, was not significantly affected, suggesting that the common element linking these effects may be a state of functional iron deficiency.

Few studies in the literature have investigated changes in EEG in cancer patients undergoing chemotherapy, and how, if at all, those changes might be related to changes in HbO. Kreukels et al. recorded EEG in breast cancer patients treated with and without various chemotherapy using an auditory oddball paradigm [71]. Their cross-sectional comparison revealed lower P3 amplitude in the chemotherapy group than the control patient group without chemotherapy, though this pattern of results was complicated by the fact that the chemotherapy patients were at more advanced disease stages. Importantly, for the first time, our longitudinal investigation using EEG in ovarian cancer patients demonstrated that the hemodynamic *but not the electrical* response, changed after chemotherapy. NVC is the process by which neuronal activity in the brain leads to localized changes in blood flow, ensuring that active regions receive adequate oxygen and nutrients by way of Hb and red blood cells. This relationship is crucial for maintaining cerebral blood flow and sufficient oxygenation, as it allows the brain to dynamically regulate perfusion based on metabolic demand. Our finding of altered NVC relates to findings of changes in CBF following chemotherapy [72–76]. Noteworthy, we were able to do this using simultaneous EEG-fNIRS recording and a simple and quick motor task, which was easily administered in the clinic setting and that demonstrated test-retest reliability and sensitivity to detect longitudinal changes.

Ovarian cancer is one of the most common gynecological malignancies. Due to the characteristics of the disease, treatment, and related symptoms, including high recurrence, extensive tumor reduction, multiple rounds of chemotherapy, and surgical menopause, nearly 69 % of ovarian cancer patients report suffering from cognitive dysfunction, which is higher than previous reports in breast cancer patients [3, 77]. The cause of the chemotherapy related cognitive impairment is thought of to be related to the increased levels of cytokines in vivo, which can lead to neurotoxicity, glial cell changes, decreased nerve repair, and accelerated aging / oxidative stress. In addition, the resultant inflammation can result in a sequestering available iron, resulting in functional iron deficiency in which iron not available for metabolic processes, including erythropoiesis.

There are quite likely multiple factors causing cognitive impairment in cancer treatment, and our unique combination of blood, hemodynamic, and electrical measurements in the context of behavioral measurements of attentional function allow for more complex relationships to be examined. Firstly, findings from our study strongly suggest a vascular effect rather than an electrical effect which may be attributed to the restricted cerebral flow resulting from reductions in red blood cells due to impaired erythropoiesis resulting from sequestered iron due to inflammation induced by the cytotoxic agents [78–80]. These results point to the importance of using HbO as a useful biomarker in addition to a range of blood iron biomarkers, in order to better elucidate the potential role of functional iron deficiency in CRCI. Additionally, our finding that the EEG ERP was not changed by the time of finishing 2-6 rounds of chemo suggests that the effects of chemotherapy on cortical oxygenation and the mass action of cortical neurons and circulations are separable. Note, however, that this does not rule out effects of functional iron deficiency on subcortical neurons, as iron deficiency has been shown in animal models to preferentially affect signaling and neurotransmitter synthesis and regulation neurons in in mid-brain regions [31, 81, 82], including basal ganglia, and the activity of such populations cannot be measured using scalp EEG. Given that the effects at the levels of cerebral HbO and systemic Hb were so strong, particularly with respect to the observed changes in attentional performance, this study indicates the potential for fNIRS to serve as an additional tool for measuring the brain’s response to chemotherapy. With further development, it could even provide a mechanism to protect cognitive function by identifying early vascular changes and intervening before more severe cognitive deficits arise [78, 80, 83].

While this study has resulted in several new findings, there are limitations. The most notable are that the final sample size was quite small and was lacking in diversity, and increasing the sample size could allow a systematic evaluation on the effect of chemotherapy on the set of measures used here in a more representative sample with greater statistical power. Another limitation of this study was a lack of whole-brain fNIRS data acquisition. While EEG was spaced evenly throughout the head, fNIRS was only placed over the left motor region. Extending the coverage to regions above the frontal and parietal cortexes could image the brain areas that underlie the distributed attentional networks that were directly engaged by the ANT. Finally, the study used only one measure of cognitive performance, and future work needs to consider other aspects of cognition relevant to subjective “chemo-fog,” such as memory, processing speed, and alertness.

In summary, this is a study that utilized simultaneous fNIRS and EEG to investigate the neurological mechanism of chemotherapy related cognitive impairment in ovarian cancer patients. This study provides compelling evidence that, in the context of treatment for ovarian cancer, chemotherapy induces significant changes in attentional performance that are accompanied by (a) significant reductions in cortical oxygen transport capability, as uniquely measured here using fNIRS; with (b) no changes in cortical electrical activity, as measured using concurrent EEG; (c) significant changes in blood iron biomarkers related to oxygen transport, particularly Hb; and with (d) no significant changes in biomarkers indicative of iron stores, particularly sFt. The significant dose-response relationship involving the fNIRS HbO responses suggests that the neurovascular coupling metrics available from concurrent fNIRS-EEG measurement could serve as a potential biomarker for detecting and monitoring cognitive impairment in cancer patients, and the multimodal imaging may be a useful tool for assessing brain vulnerability to the effects of chemotherapy. Additionally, the association between cognitive functions and systematic Hb levels, in the context of no significant change in sFt, point to the possibility of emerging effects of functional iron deficiency, systematically and within the brain, the latter of which was uniquely measurable using our fNIRS-EEG approach. Our findings suggest avenues for early intervention strategies aimed at protecting cognitive function during and after cancer treatment, as well as improved chemotherapy developments.

## Methods

### Participants

All study protocols were approved by the Institutional Review Boards at The University of Oklahoma Health Sciences Center (OUHSC) and the patient protocol was also approved by the OUHSC Stephenson Cancer Center. All procedures involving the research participants have been performed in accordance with the Declaration of Helsinki.

Patients aged 19-99 years with stage III-IV epithelial ovarian or endometrial cancer were recruited and signed written consent before any experimental procedures. The inclusion criteria considered both ovarian and endometrial cancer patients, as they are typically treated with a common protocol and present very similar neurocognitive impairment [59, 60]. The patients were screened to exclude persons with existing neurological or neuropsychiatric diseases. All subjects had to be fluent in English, have normal or corrected to normal vision and hearing, and unimpeded use of both hands.

In parallel to the patient study, healthy participants aged between 18 and 60 were recruited and studied to assess the reliability of the neurovascular coupling protocol. Since the healthy subjects were studied to primarily evaluate the reliability of the NVC measures, they were not age matched to the cancer patients. Informed consent forms were signed before any experimental procedures. Subjects were screened to exclude persons with existing neurological or neuropsychiatric disorders. All subjects had to be fluent in English and have normal or corrected to normal vision and hearing.

### Experimental Design

For the cancer patients, there were two repeated visits in the study design: one visit at diagnosis and before chemotherapy or within the initial round of chemotherapy, and one visit after at least three rounds of chemotherapy. Each visit consisted of acquisition of a structural magnetic resonance image (MRI), standard assays for blood iron biomarkers, and cognitive testing with concurrent fNIRS-EEG recording. The MRI results are not discussed in this paper. fNIRS-EEG cognitive testing consisted of five tasks in this order: motor 1, motor 2, resting state 1, ANT, and resting state 2. Each task was custom-coded using E-Prime 2.0 (Psychology Software Tools, Pittsburgh PA). During resting state tasks, subjects fixated on a cross at the center of a laptop screen for 6 min; they were instructed to keep their eyes open during rest. During the motor tasks, subjects performed 7 blocks of hand clenching at the frequency of 1 Hz. An initial cross would appear for 30 s, then a 1 Hz blinking arrow would appear for 20 s guiding fist clenching, followed by a 30-s resting block; this pattern repeated for 6 min. Subjects clenched their right hand at 1 Hz with the visual cue.

The ANT consisted of 180 trials in which participants judged the direction with which an arrow at the center of screen was pointing. Trials began with a fixation cross, followed by a response cue and then the test stimulus display. The response cue was either no cues, two cues (one below and one above fixation), or a valid spatial cue, either above or below fixation. The test stimulus display contained five elements, a central arrow pointing either left or right, flanked by two elements on the left and two elements on the right. These elements could either be neutral (no arrowheads), consistent with the central arrow (arrows pointing the same direction), or inconsistent with the central arrow (arrows pointing the opposite direction). Participants made their responses using the ‘z’ (for left) and ‘m’ (for right) keys on the computer keyboard. Both response accuracy and RT were recorded on each trial, and the RT difference scores (described below) were calculated using only correct responses.

For the healthy subject study, two repeated visits occurred within 14 days of one-another. This time frame ensured there would not be drastically different neural responses in healthy subjects. Each visit consisted of fNIRS-EEG recordings during six tasks: resting 1, motor 1, motor 2, auditory 1, auditory 2, resting 2. Auditory recordings are not discussed in this paper. The same motor and resting tasks were used as in the cancer-subject design. Healthy subjects did not have MRI, bloodwork or cognitive testing.

### Data Acquisition

For both cancer and healthy participants, simultaneous fNIRS, EEG, and peripheral measurements of pulse oximetry, accelerometry, and respiration were recorded in every subject, following our prior recording protocol [47, 54].

In the cancer patient study, fNIRS measurements were acquired using a NIRScout system (NIRx Medical Technologies, LLC, New York, United States). Eight source probes, seven detector probes, and eight short-separation detectors were arranged over the left motor region totaling 30 channels (i.e. 30 pairs of sources and detectors). Among these, eight short-separation channels were evenly spread throughout. Signals were acquired at a sampling rate of 7.81 Hz. EEG measurements were acquired by a Brain Vision actiCHamp 64 channel, fNIRS compatible system (BrainProducts, München, Germany). A total of 32 EEG channels were spaced evenly across the head. The electrode at Cz was selected as the reference while ground was placed at FP1. Electrically conductive gel was used and impedances of the EEG electrodes were maintained throughout under 20 kΩ. EEG data was acquired at a sampling rate of 500 Hz.

In the healthy participant study, fNIRS measurements were acquired by a NIRScout system (NIRx Medical Technologies, LLC, New York, United States). A total of 16 source probes, 26 detector probes, and 16 short-separation detectors were arranged over the bilateral motor and auditory regions, totaling 76 channels. Of these, 16 short-separation channels were evenly spread throughout. The fNIRS data was collected at a sampling rate of 3.91 Hz. EEG measurements were acquired by a Brain Vision actiCHamp 64 channel, fNIRS compatible system (BrainProducts, München, Germany). A total of 32 EEG channels were evenly placed throughout the head. The electrode at Cz position was selected as the reference while ground was placed at FP1. Electrically conductive gel was used and impedances of the EEG electrodes were maintained at under 20 kΩ throughout. EEG data was acquired at a sampling rate of 500 Hz.

### Laboratory Tests

Participants underwent a blood draw to determine iron status. Blood samples were obtained using antecubital venipuncture by trained phlebotomists at the University of Oklahoma Health Sciences Center. Laboratory tests were performed on these samples in the laboratories of OU Medicine using standard clinical practices. The variables of interest were hemoglobin (Hb, g/dL), serum ferritin (sFt, ng/mL), mean corpuscular volume (MCV, fL), hematocrit (HCT, %), mean corpuscular hemoglobin (MCH, pg), mean corpuscular hemoglobin concentration (MCHC, g/dL), red blood cell count (RBC), red blood cell distribution width (RDW, %), and C-reactive protein (CRP, mg/L), with the latter being used to assess inflammation.

### Behavioral Data Analysis

Consistent with the design of the ANT as an RT task, accuracies were consistently near ceiling for all participants at both assessments and so will not be discussed. Median RTs for correct responses were calculated for each participant in each of the combinations of cueing and flanker conditions, and these medians were used to calculate three difference scores. In each case, higher values are indicative of better attentional function. The first of the difference scores indexes the extent to which low level attention can be captured by the onset of stimulus energy and is the difference between RT in the 0-cue condition and RT in the two-cue condition; this is referred to as the alerting score. The second of the difference scores indexes the extent to which the participant can use a spatial cue to orient attention in advance of the test display and is the difference between the RT in the central cue condition and the RT in the (valid) spatial cue condition; this is referred to as the orienting score. The third difference score indexes the extent to which a participant is capable of benefitting from consistent elements of the stimulus and avoiding costs from inconsistent elements and is the difference between the RT in the inconsistent flanker condition and the RT in the consistent flanker condition; this is referred to as the conflict score. All analyses were done using SAS 9.4 for Linux and R 4.4.1.

### fNIRS Data Analysis

The following preprocessing steps were performed on the data from cancer and healthy subjects, following our prior protocol for the PCA-GLM pipeline [47, 54]. Physiological noise due to superficial tissue absorption, respiration, cardiac pulsation and head motion were removed, resulting in relative changes of Hb and HbR. Prior studies of ours and other groups have indicated that fNIRS HbO has higher sensitivity than HbR; considering our primary objective in the study is to detect chemotherapy related changes, our analysis focused on the HbO. Then, a 5-channel region of interest (ROI) over the motor cortex was chosen for targeted analysis. Block averages with reference to the beginning of clenching blocks were calculated. To quantify the task-related responses, the time courses of the block average was further averaged between 5-20 s, considering the delay of the hemodynamic response. These values were exported as HbO and HbR values for fNIRS and NVC comparisons. All analyses were done using customized scripts in MATLAB^®^ 2023a.

### EEG Data Analysis

The following preprocessing steps were performed on the EEG data from the cancer patients and the healthy subjects, following our prior protocol [47, 54]. The EEGLAB toolbox [61] and customized scripts in MATLAB^®^ 2023a were used. Once raw data was loaded to the toolbox with channel locations, it was filtered with bandpass and notch filters, extracted into epochs relative to the onset cues, and ocular and muscular noise was removed with EEGLAB’s independent component analysis. Epochs were visually inspected and those with excessive motion artifacts were manually rejected. Once all noise was removed, the data was re-referenced to the common average reference and averaged across all epochs, resulting in the event-related potentials (ERP).

### Test-retest Reliability Assessment

In parallel to studying NVC in cancer patients, reliability was assessed in healthy subjects with two visits. Identical processing steps of EEG and fNIRS were performed in the healthy subjects’ data from each visit. After quantifying the fNIRS and EEG responses, a two-tailed, paired *t*-test assuming unequal variances was applied to the test and retest in the healthy. To identify which quantity was more reliable, a one-way random effect ICC was calculated [62]. For these measurements, the primary components are the between-subject variance that uses between-subjects mean squares (MS_B_) and the between-test variance that uses within-subject mean squares (MS_W_). The following equation was used to quantify the reliability:

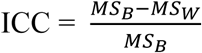

Reliability was determined using a 0.0 to 1.0 scale that ranged from poor (ICC < 0.40), fair (0.40 < ICC < 0.59), good (0.60 < ICC < 0.74) to excellent (0.75 < ICC < 1.00) [62].

## Author Contributions

SE: Investigation, Formal analysis, Methodology, Writing - original draft. QS: Investigation, Formal analysis, Methodology, Writing - original draft. FZ: Investigation, Methodology, Writing - original draft. AKC: Investigation, Writing - original draft. LD: Writing - original draft. DMB: Conceptualization, Investigation, Writing - original draft. JW: Conceptualization, Investigation, Writing - original draft. DW: Investigation, Writing - original draft. MW: Conceptualization, Methodology, Formal analysis, Funding acquisition, Writing - original draft, Supervision. HY: Conceptualization, Methodology, Formal analysis, Funding acquisition, Supervision, Writing - original draft.

## Data Availability Statement

Data used in this study are not publicly available due to data sharing restrictions from the IRB but are available from the corresponding author through a data use agreement upon reasonable request.

## Acknowledgements

We thank the Clinical Trials Office of the Stephenson Cancer Center at University of Oklahoma Health Sciences Center for their collaboration and support.

